# Efficacy of Connexin testing in detecting Non-syndromic Sensorineural Hearing Loss in an Indian cohort

**DOI:** 10.1101/2020.07.30.20165530

**Authors:** Jagannath Kurva, Nalini Bhat, Suresh Shettigar, Harshada Tawade, Shagufta Shaikh, Anu Ghosh, Susan Cherian

**Affiliations:** Cytogenetics and Molecular Genetics Section, Pathology Unit, BARC Hospital, Medical Division, Bhabha Atomic Research Centre (BARC), Anushakti Nagar, Mumbai-400094, India; ENT Unit, BARC Hospital, Medical Division, Bhabha Atomic Research Centre, Anushakti Nagar, Mumbai-400094, India; Radiation Biology & Health Sciences Division, Bhabha Atomic Research Centre, Trombay, Mumbai-400085, India; Homi Bhabha National Institute (HBNI), Anushakti Nagar, Mumbai-400094, India

**Author notes:** **Corresponding author**, E-mail address, Full Postal Address: Cytogenetics and Molecular Genetics Section, Pathology Unit, BARC Hospital, Medical Division, Bhabha Atomic Research Centre, Anushakti Nagar, Mumbai-400094, India.

**Keywords:** Non-syndromic Sensorineural Hearing Loss, *GJB2*, *GJB3*, *GJB6*

## Abstract

Hearing loss is one of the most common sensory disorder and approximately 466 million people have disabling hearing loss worldwide. This study was conducted to identify the mutations in the *GJB2, GJB3*, and *GJB6* genes in an Indian cohort with non-syndromic sensorineural hearing loss and ascertain its use for genetic testing. 31 affected individuals with prelingual bilateral non-syndromic severe to profound sensorineural hearing loss were identified based on clinical evaluation and audiometric assessment. Sanger Sequencing method was used. Six out of 31 affected individuals showed pathogenic nonsense mutations in *GJB2* gene, accounting to 19.3%. Of the 6 affected individuals, 5 were homozygous for c.71G>A(p.Trp24Ter) and one was compound heterozygous for c.71G>A and c.370C>T(p.Gln124Ter). Missense mutations [c.380G>A(p.Arg127His) and c.457G>A(p.Val153Ile)], and 3’ UTR variations were also identified in *GJB2* gene. *GJB3* and *GJB6* genes showed only silent mutations and 3’ UTR variations. 19.3% of affected individuals showing pathogenic mutations in *GJB2* gene in our cohort is comparable to other Indian studies (approximately 20%) and it is less as compared to Caucasian, Japanese, and Chinese studies (approximately 50%). Lower occurrence of pathogenic mutations in *GJB2* gene in our cohort and other Indian studies as compared to other Caucasian, Japanese and Chinese studies, and absence of pathogenic mutations in *GJB3* and *GJB6* genes indicates that these genes may have a limited role in the Indian population. Hence there is a need to identify genes that play a major role in the Indian population so that they can be used for genetic testing for NHSL to aid in accurate and early diagnosis.

## INTRODUCTION

Hearing loss (HL) is one of the most common sensory disorder and approximately 466 million people have disabling hearing loss worldwide [1]. HL in infants leads to serious problems in speech and education and impairs the quality of life. Approximately 50- 60% of the HL is due to genetic causes [2]. Till date, mutations in approximately 180 genes are associated with HL. Of these 180 genes, 124 are associated with non-syndromic hearing loss (NSHL) (http://hereditaryhearingloss.org/). Of these 124, *GJB2* is the most common gene associated with NSHL and has a high frequency of variations, especially in its coding sequence (ENSG00000165474). Owing to this, *GJB2* mutation testing has become an important tool for diagnosis [3]. According to various studies, approximately 50% of NSHL is caused by mutations in the *GJB2* gene in Caucasian, Japanese, Chinese, and the Ashkenazi Jewish population [4–7]. It is also known, mutations in *GJB2* and *GJB3* genes can cause NSHL in a digenic pattern of inheritance [8]. Heterozygous mutations in *GJB2* gene and a concurrent deletion in *GJB6* gene can also contribute to NSHL in a digenic pattern of inheritance. A 342-kb deletion is the most common mutation in the *GJB6* gene in Caucasian studies [9, 10].

The current world wide genetic testing protocol for NSHL as recommended by the ACMG involves the first step of screening GJB2, GJB6 for mutations and deletions[11]. Our cohort of approximately 100000 individuals is a stable population derived from the employees of Department of Atomic Energy and their immediate family members. This is fairly representative of the Indian population, although there is a predominance of local population of Maharashtra state of India.

This study was designed to test the efficacy of first line of genetic testing used worldwide to identify the cause of NSHL in our cohort. We tested for variants in *GJB2, GJB3* and *GJB6* genes from our NSHL patients with congenital prelingual bilateral non-syndromic severe to profound sensorineural HL. We included GJB3 as it is known to cause NSHL in a digenic pattern of inheritance. Variant analysis of these genes were carried out in 31 affected individuals and it was found that 19.3% (6/31) of affected individuals had pathogenic nonsense variants in the *GJB2* gene. *GJB2* gene also showed missense variants and 3’ UTR variations. *GJB3* and *GJB6* genes were identified to have silent variants and 3’ UTR variations.

## MATERIALS AND METHODS

### CLINICAL EVALUATION

Patients (all age groups) with congenital prelingual bilateral non-syndromic severe to profound sensorineural HL (affected individuals) were identified from the New-born Screening Programme and also the subjects visiting the ENT unit of Bhabha Atomic Research Centre Hospital. Our Centre is located in western India. Although the profile of the cohort is of mixed ethnicity, there is a predominance of the native inhabitants of Maharashtra state of India. Inclusion and exclusion criteria for this study was clearly decided and used to select the subjects (Supplementary Table S1). Based on these, Clinical evaluation was carried out and etiologies other than genetic origin were screened out (Supplementary Table S2). Finally, 31 affected individuals from 25 families were selected based on the clinical evaluation and audiometric assessment. Of the 25 families, 9 were consanguineous and comprised of 10 affected individuals.

### Ethics

The study was approved by the Institutional Ethics review board and informed consent was obtained from all the individuals.

### GENETIC EVALUATION

2ml of citrate anti-coagulated peripheral blood samples were used to extract DNA using QIAamp DNA Blood Midi Kit, (Qiagen, Hilden, Germany) according to the manufacturer’s instructions. The DNA quality and quantity were assessed using spectrophotometer Nanodrop 1000.

### Primer designing

Overlapping primers were designed for all exons of *GJB2* and *GJB6* genes, and for CDS region of *GJB3* gene using ExonPrimer, IHG (http://ihg.helmholtz-muenchen.de/ihg/ExonPrimer.html) and Primer-BLAST, NCBI (https://www.ncbi.nlm.nih.gov/tools/primer-blast/). All the primers sets were checked for uniqueness and accuracy using Primer-BLAST (NCBI). PCR conditions were standardized to generate a single clean product. All the products so obtained were evaluated on DNA screening gel using the Qiaxcel Advanced system (Qiagen, Hilden, Germany). The PCR primers and amplification conditions are mentioned in Supplementary Table S3.

### Sequencing and analysis

All the PCR products were sequenced (Sanger Sequencing method) bidirectionally using ABI 3730xl DNA Analyzer and BigDye Terminator v3.1 Sequencing Kit. The ABI sequencing files were analyzed using BioEdit 7.2.5 [12] and CodonCode Aligner ver. 7.1.2. (Trial). Sequences were aligned to both, the reference sequence from RefSeqGene, NCBI (https://www.ncbi.nlm.nih.gov/refseq/) and from Ensembl Genome Browser with known variants (https://asia.ensembl.org/index.html). The identified variations were checked with the dbSNP database (http://www.ncbi.nlm.nih.gov/snp/), Clinvar database (https://www.ncbi.nlm.nih.gov/clinvar/) and the 1000 genomes database (http://www.1000genomes.org/) for pathogenicity and allele frequencies.

## RESULTS

A total of 16 variations were identified in 31 affected individuals (Table 1). Of the 16 variations, 9 were identified in *GJB2* gene, 4 variations in *GJB3* gene and 3 variations in *GJB6* gene. Only *GJB2* showed pathogenic nonsense variants and other missense variants whereas silent variants and UTR variations were seen in all the three genes.

**Table 1:**
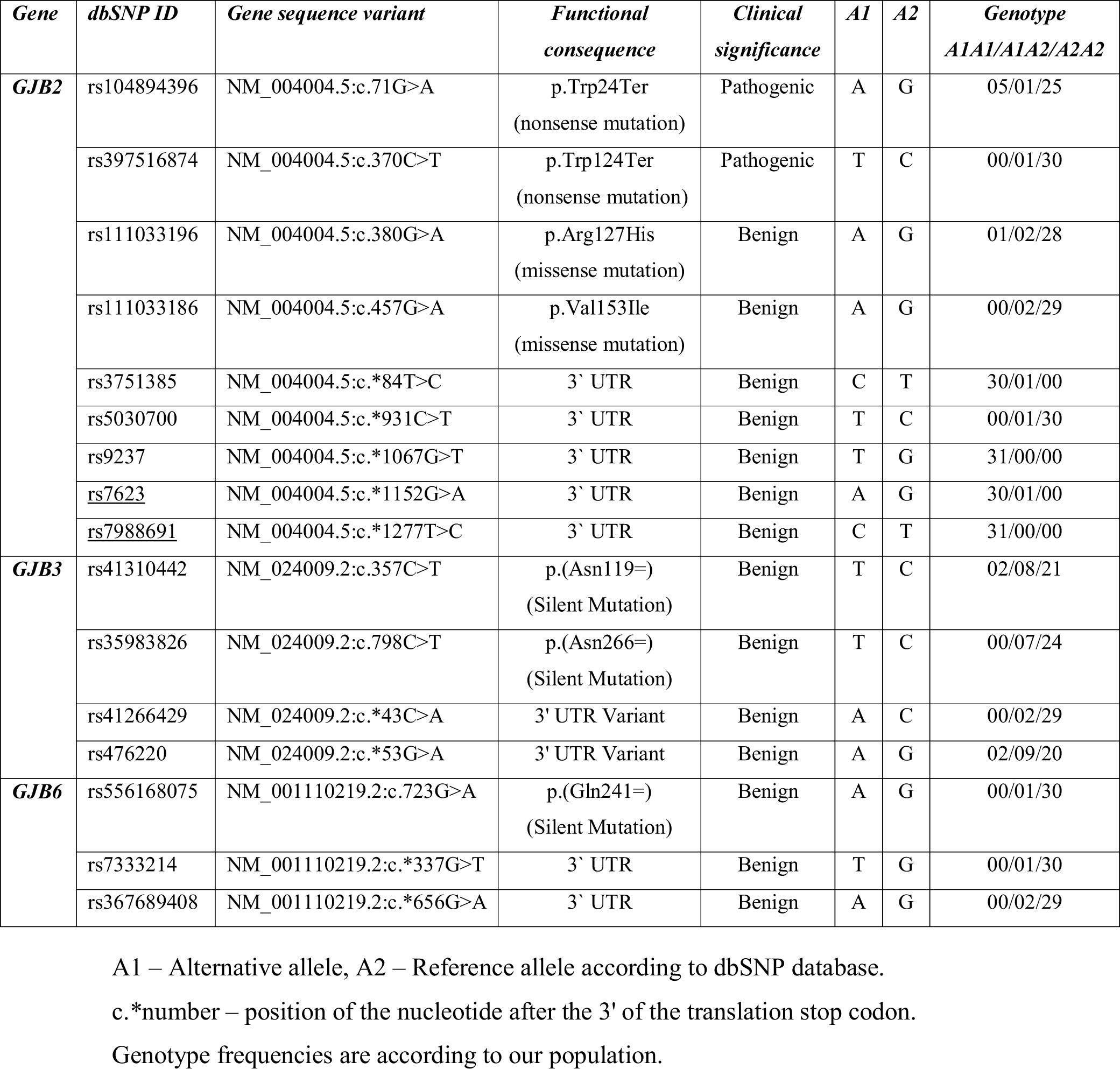
Genetic variations identified in affected individuals (n=31)

### *GJB2* GENE

A total of 9 different variants were identified in *GJB2* gene, of which 2 variants were nonsense variants, 2 were missense variants and 5 variants were seen in the 3’ UTR region.

### Nonsense Variants

Six affected individuals were identified with nonsense variants, of which 5 were homozygous for c.71G>A(p.Trp24Ter) while one was a compound heterozygous for c.71G>A and c.370C>T(p.Gln124Ter).

### Missense Variants

Two missense variants were identified in five affected individuals. Of the five affected individuals, one was homozygous and two were heterozygous for c.380G>A(p.Arg127His) and the remaining two were heterozygous for c.457G>A(p.Val153Ile) variant.

### 3’ UTR variations

Five different 3’ UTR variations were identified in *GJB2* gene in our cohort.

### Pedigree Analysis

To study the inheritance pattern of three coding variants c.71G>A, c.370C>T, and c.380G>A, all the members from the five families (Family 3, Family 5, Family 10, Family 15 and Family 24) were Sanger sequenced and their pedigrees were analyzed (Figure 1).

**Figure 1:**
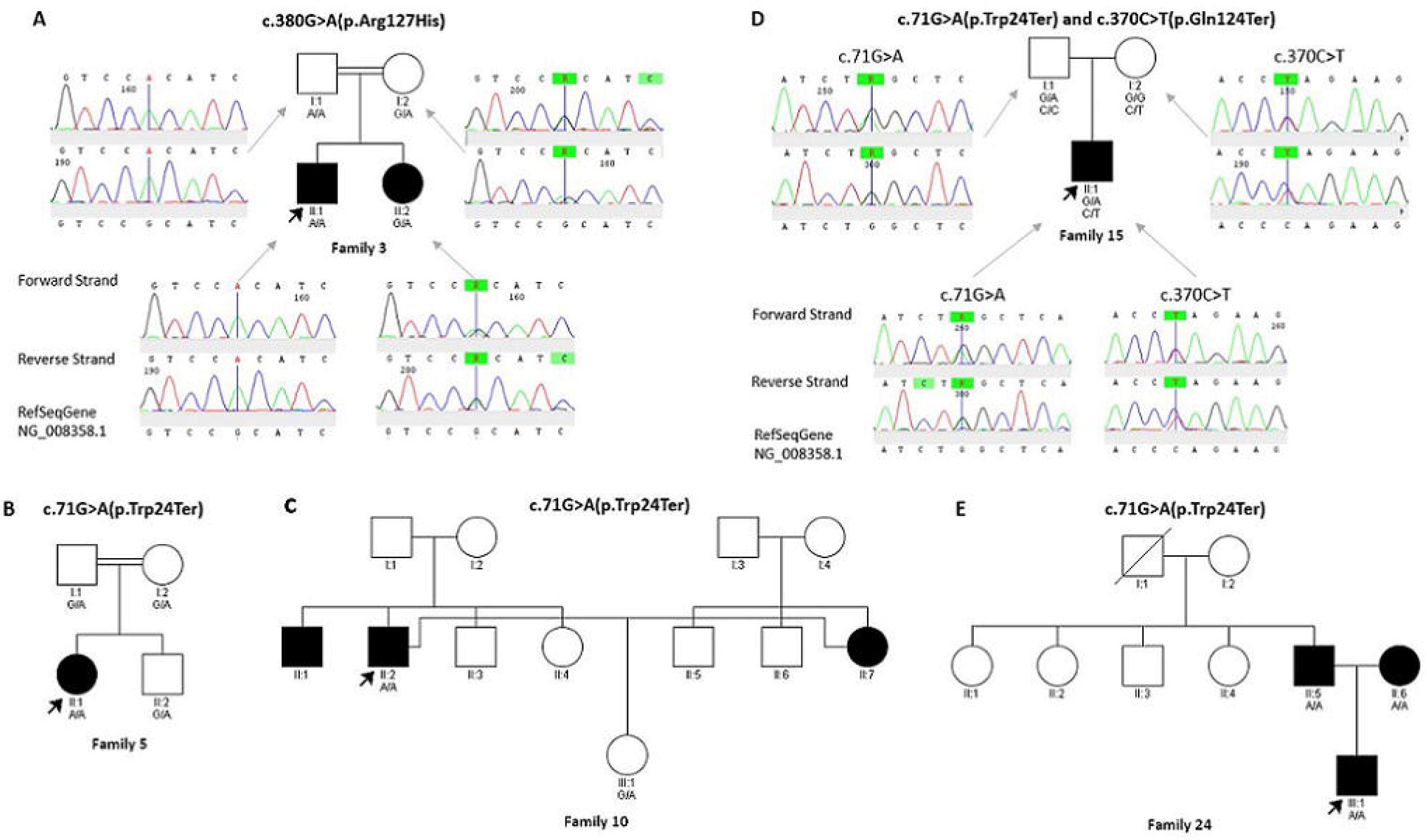
Pedigree of Family 3, Family 5, Family 10, Family 15, and Family 24. All mutations mentioned in the figure from GJB2 gene. (A) Pedigree and electropherograms showing the inheritance pattern of missense mutation c.380G>A in Family 3. (B) Pedigree showing the inheritance pattern of nonsense mutation c.71G>A in Family 5.(C) Pedigree depicting the inheritance pattern of nonsense mutation c.71G>A(p.Trp24Ter) in Family 10. (D) Pedigree of Family 15 shows inheritance pattern of compound heterozygote of c.71G>A and c.370C>T along with electropherograms. (E) Pedigree showing the inheritance pattern of nonsense mutation c.71G>A in Family 24.

The c.71G>A and c.370C>T variants are known to be pathogenic and inherited in homozygous recessive pattern. In Family 3, the proband was homozygous and his affected sibling was heterozygous for c.380G>A, whereas their unaffected father was also homozygous and their unaffected mother was heterozygous for the same c.380G>A variant. The variant c.380G>A is a benign variant rather than a pathogenic variant and the causal gene in this family must be different than the selected genes.. In Family 5, the proband was homozygous, while parents were heterozygous for c.71G>A variant. In case of Family 10, the proband is homozygous for c.71G>A, and his wife also affected with NHSL, did not show any pathogenic variants in the selected genes for this study indicates that there could be other genes playing role in HL. Their offspring is normal and was found to be a carrier for c.71G>A mutation. Family 15 is the best example of the compound heterozygous pattern of inheritance. Proband is a compound heterozygous for c.71G>A and c.370C>T variants. Proband inherited these variants from each heterozygous parents. In Family 24, the proband and both his affected parents were found to be homozygous for c.71G>A pathogenic variant. The proband had a cochlear implant and hence is able to communicate verbally due to newborn screening and subsequent intervention.

### *GJB3* GENE

Four variations were identified, two of which were silent variants and other two were in the 3’ UTR region. All variants were benign in nature.

### *GJB6* GENE

Three variations were identified in the *GJB6* gene, of which one was a synonymous variant while the other two variations were in the 3’ UTR region. We did not find any pathogenic variants in *GJB6* gene which is consistent with other reports from Indian studies [13–15].

## DISCUSSION

In *GJB2* gene mutations, c.71G>A in our cohort was found to be the most common pathogenic variant and identified in six of the 31 affected individuals. Some authors consider it to be the predominant pathogenic variant causing NSHL in India [16, 17]. This dominance of c.71G>A in Indian population could be a result of founder effect [16].

Clinical significance of c.380G>A and c.457G>A variants are categorized as “OTHERS” type because protein function prediction softwares like SIFT, Polyphen2 found these variations as benign, while Mutation taster and PANTHER classified it as pathogenic [18]. Our frequency distribution study(data not shown) and pedigree analysis reaffirms c.380G>A is not a pathogenic variant and does not cause the condition.

All the variants were already described in the existing databases. The commonly found *GJB2* mutation c.35delG among Whites and c.235delC among Chinese and Japanese were not seen in our cohort [6, 19–22], although the variant c.35delG was reported in another study on the North Indian population [23]. In addition, we did not find any digenic heterozygote mutations in our cohort.

We found 2 synonymous variants and 2 UTR variants in *GJB3* in our cohort. Some studies have suggested that *GJB3* may be an uncommon cause of NHSL [24–26]. We found 1 synonymous variant and 2 UTR variants in *GJB6* gene. It is commonly known that 342kb deletion in GJB6 gene along with mutations in *GJB2* gene can cause NSHL in digenic pattern of inheritance [9, 27].

## CONCLUSION

Our genetic testing for pathogenic variants in the selected connexin genes could identify the cause of NSHL in only 19.3% (6/31) of our cohort. The low frequency of pathogenic variants detected in *GJB2* gene is not comparable to Caucasian, Japanese and Chinese population whose frequency is between 30-50%. None of Indian studies including current study have found the deletion in GJB6 gene, which acts in a digenic pattern of inheritance in other world populations. Pathogenic variants in Indian population, are different to ones normally seen in other world populations. This has implications in using certain genetic tests recommended by regulatory authorities in other population. This will lead to firms providing genetic test to only include common variants from world population without representation from Indian population. Owing to the above inferences, it can be said that genetic testing of only *GJB2, GJB3*, and *GJB6* have a limited role in the Indian population. Although the current first level testing would be able to identify the cause in 1/5th of NSHL patients, there is a need to identify additional genetic markers for an accurate and early genetic diagnosis in the Indian population. A whole exome or whole genome approach to detect the genetic markers in Indian NSHL cohorts, would help in achieving a better first level genetic tests for NSHL.

## Data Availability

All the important data is included in the manuscript and additional data is provided in the supplementary data

## ACKNOWLEDGEMENTS

Our sincere gratitude to Dr. M Seshadri for his critical inputs during the study and during the writing of the manuscript. We also thank Ms. Shailaja Chavali for the collection of samples and DNA extraction of a few samples.

## SUPPLEMENTARY INFORMATION

Supplementary Table S1 Supplementary Table S2 Supplementary Table S3

